# Retinal perfusion delay associated with GLP-1 receptor agonists: A possible role of intracranial pressure?

**DOI:** 10.1101/2025.09.25.25336128

**Authors:** Yun Kyung Lee, Yuan Yi Chang, Han Na Jang, Dae Joong Ma, Hyun Beom Song

**Author notes:** Corresponding author: Hyun Beom Song, MD, PhD, Department of Biomedical Sciences, Ophthalmology, Tropical Medicine and Parasitology, Seoul National University College of Medicine, 103 Dahakro, Jongnogu, Seoul, Republic of Korea.

## Abstract

Recent studies have suggested an association between GLP-1 receptor agonist (GLP-1RA) treatment and increased risk of non-arteritic anterior ischemic optic neuropathy, particularly within the first year of treatment. This study investigated potential mechanisms by examining retinal perfusion changes in both humans and mice during GLP-1RA therapy. We first compared six patients who underwent fluorescein angiography during GLP-1RA treatment with three patients after treatment completion. Two cases in a during-treatment group showed increased arteriovenous transit time. To validate these findings, we treated wild-type mice with liraglutide and measured retinal perfusion parameters before, during, and after treatment. Mice exhibited delayed injection-to-retina time and increased arteriovenous transit time during treatment, and decreased scotopic b-wave responses and increased hypoxia-related gene expression indicating retinal hypoxia, with recovery after treatment discontinuation. Investigation of mechanisms revealed that GLP-1RA-treated mice had significantly reduced intracranial pressure (ICP) at day 7, corresponding with the timing of retinal perfusion delays, and there was a trend toward a negative correlation between ICP and injection-to-retina time. Reduced ICP may affect ophthalmic vessels in the subarachnoid space, contributing to delayed retinal perfusion. These findings suggest that ocular circulation may be particularly vulnerable to GLP-1RA-related hemodynamic effects, independent of systemic cardiovascular changes. These results indicate that careful monitoring of ocular circulation may be warranted in individuals using GLP-1RA, and suggest the need for further investigation of temporal ICP changes in human patients.

## Main

In recent years, glucagon-like peptide-1 receptor agonists (GLP-1RAs) have received significant attention in the medical field. GLP-1RA selectively binds and activates GLP-1R mimicking the effect of glucagon-like peptide-1 (GLP-1), which plays a pivotal role in regulating blood glucose levels and lipid metabolism. In addition to the approved indication for type 2 diabetes mellitus (T2DM) and obesity, GLP-1RAs have been shown to have favorable hemodynamic effects, reducing blood pressure in patients with diabetes and pulmonary artery pressure in patients with heart failure^1-3^. However, recent retrospective observational studies have suggested a potential association between semaglutide treatment and an increased risk of non-arteritic anterior ischemic optic neuropathy (NAION)^4-10^. The risk was highest within the first year of semaglutide prescription according to the survival analyses in population with T2DM and overweight/obesity^5^. Even in individuals without these underlying conditions, GLP-1RA, particularly liraglutide, was associated with an increased risk of NAION within the first year of treatment^11^. These findings suggest that certain risk groups for NAION may exist independent of comorbidity status, and that subthreshold hemodynamic changes limited to ocular circulation might be detectable in individuals who have not developed NAION.

First, we identified 15 patients who underwent fluorescein angiography (FAG) before and after GLP-1RA prescription between 2016 and 2024. Among them, nine had matched series that allowed evaluation of the arteriovenous transit time (AVTT) in the same eye. We compared six patients who underwent FAG during GLP-1RA treatment with three patients who underwent FAG after completion of the treatment. Although the sample size was limited, basic demographic variables, including body weight, BMI, HbA1c and diabetic retinopathy grading, did not differ significantly between the two groups **(Extended Table 1)**. Interestingly, we identified two cases with increased AVTT (the interval between arterial filling and the venous phase) on FAG, all of which were in the group that underwent FAG during GLP-1RA treatment and all of which showed progression of diabetic retinopathy from moderate nonproliferative diabetic retinopathy (NPDR) to severe NPDR **(Fig. 1a, b)**. Although we could not identify specific risk factors for increased AVTT in this dataset, these findings suggest that a subgroup of individuals under GLP-1RA treatment exhibited increased AVTT, whereas those after discontinuation did not.

**Figure 1.**
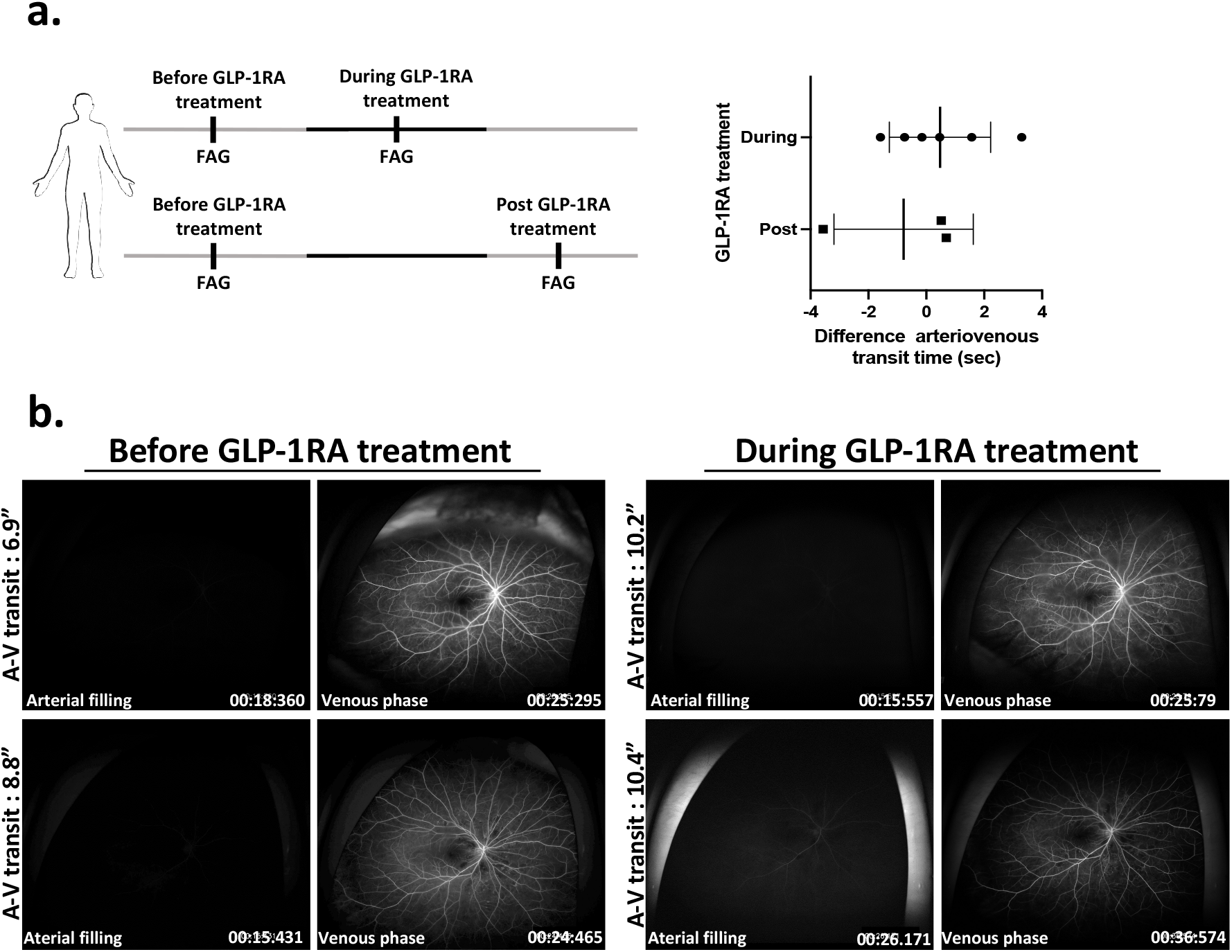
Increased arteriovenous transit time in subjects during treatment with GLP-1 receptor agonist dulaglutide. **a**. Timeline of fluorescein angiography (FAG) examinations conducted before and after GLP-1RA treatment in nine patients with type 2 diabetes, and changes in atrioventricular transit time (AVTT) observed in the during-GLP-1RA treatment group (n=6) and the post-GLP-1RA treatment group (n=3). **b**. Representative FAG images from two subjects in the during-treatment group, showing increased arteriovenous transit time following GLP-1RA administration.

Then, we tried to simulate these findings in wild-type mice by evaluating retinal perfusion after treatment with GLP-1RA, liraglutide, and measure relevant parameters both during and after discontinuation of the treatment **(Fig. 2a)**. When the same eye was followed up before and after treatment, injection-to-retina time was delayed on days 7 and 21 **(Fig. 2b, c)**, and AVTT was delayed on day 7 after treatment **(Fig. 2b, d)**. Then, the anatomical and functional impacts of delayed perfusion were evaluated using optical coherence tomography (OCT) and electroretinography (ERG), respectively. While OCT did not demonstrate any differences between the treated and control groups on day 7 **(Extended Fig. 1a)**, the GLP-1RA treated group exhibited decreased scotopic b-wave responses on days 10 and 17 after treatment **(Fig. 2e)**. Further follow-up revealed recovery of the decreased b-wave responses by day 21 after discontinuation of the treatment **(Fig. 2e)**. As hypoxia is known to reduce b-wave amplitude in humans and mice^12,13^, the expression of hypoxia-related genes, such as *Hif1*_α_, *Vegf*_α_, and *Epo*, was evaluated. Their expression was increased on day 21 after treatment but returned to normal levels by day 21 after discontinuation of treatment **(Fig. 2f)**, showing a pattern similar to the ERG responses. Immunostaining of Hif1_α_ also supported the changes induced by GLP1-RA treatment **(Fig. 2g)**.

**Figure 2.**
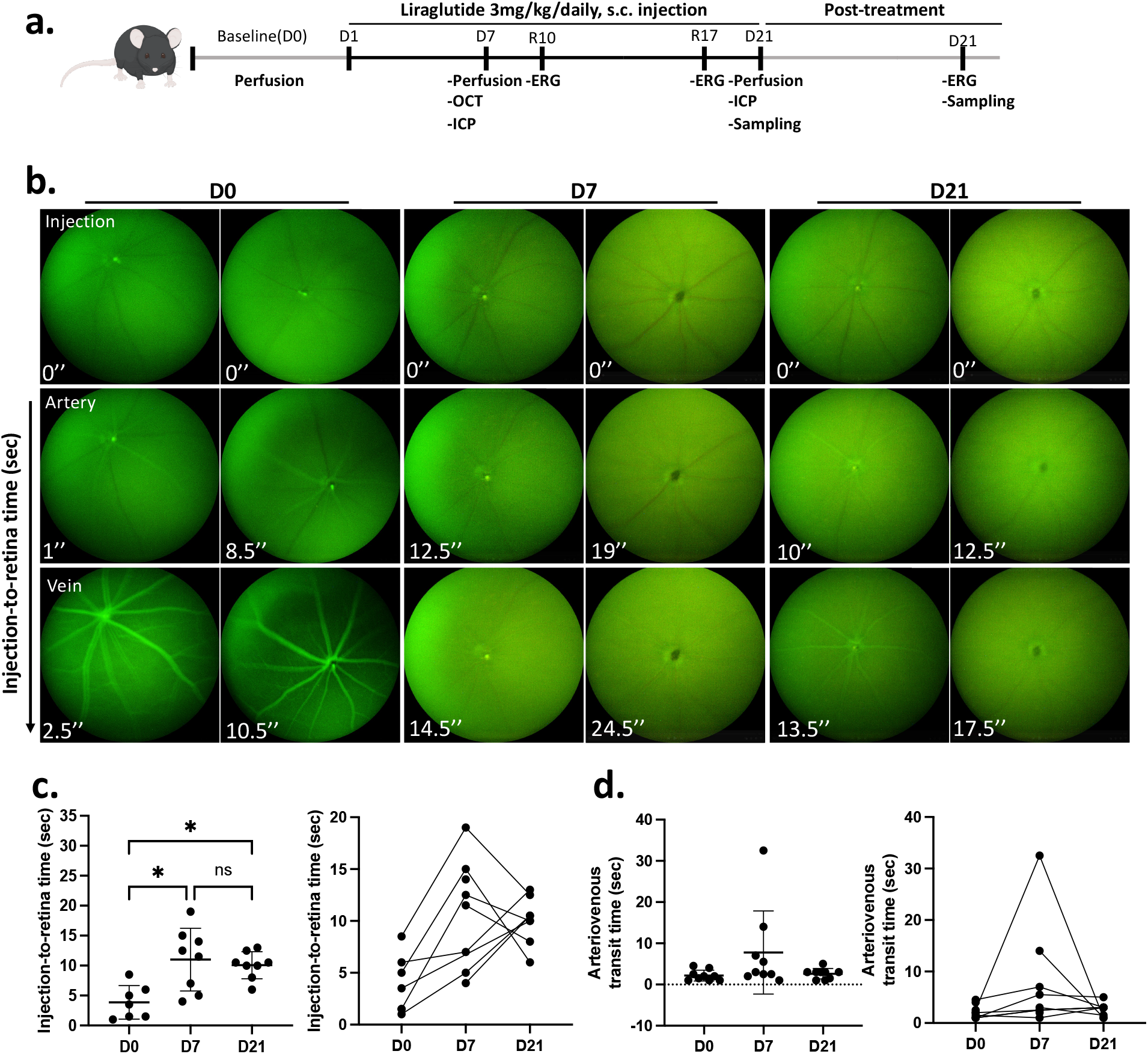
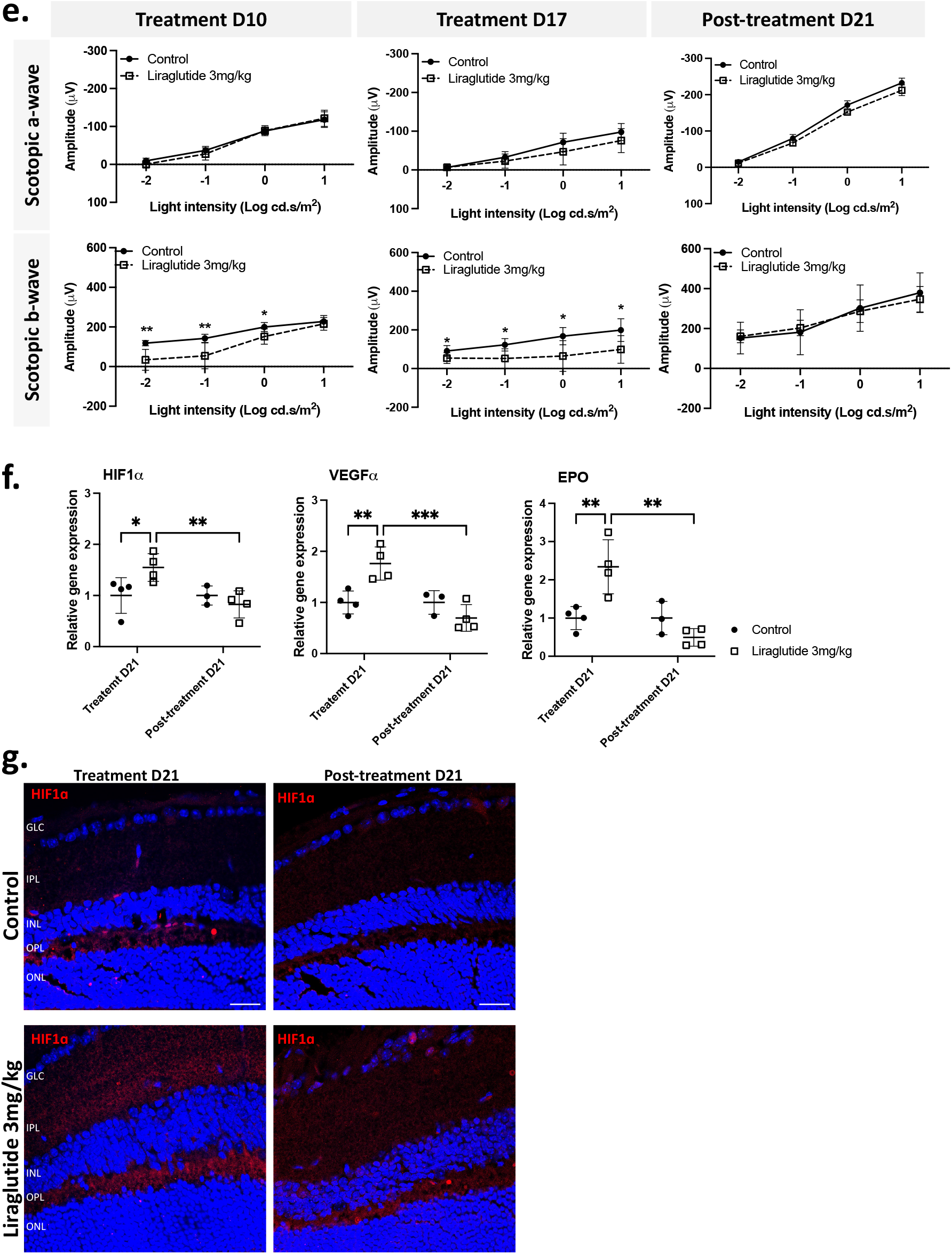
Increased injection-to-retina time and arteriovenous transit time and associated hypoxia-related changes that recover after cessation in GLP-1RA liraglutide-treated mice. **a**. Experimental flow chart. **b**. Representative longitudinal images from fundus fluorescein angiography at baseline (D0), at day 7 (D7), and at day 21 (D21) after daily administration of 3 mg/kg liraglutide in the wild-type mice (n=7). **c**. Analysis of injection-to-retina time at baseline (D0), at day 7 (D7), and at day 21 (D21) after daily liraglutide administration. **d**. Analysis of arteriovenous transit time at baseline (D0), at day 7 (D7), and at day 21 (D21) after daily liraglutide administration. **e**. Scotopic electroretinogram responses at day 10 and at day 17 after daily liraglutide administration (n=7 treated *vs*. 8 control), and at day 21 after cessation of liraglutide administration (n=3 treated *vs*. 4 control). **f**. Hypoxia-related gene expression at day 21 after daily administration of 3 mg/kg liraglutide (n=4 *vs*. 4 control), and at day 21 after cessation of liraglutide administration (n=3 treated *vs* 4 control). **g**. Hif1α protein expression at day 21 (D21) after daily liraglutide administration (n=3 *vs*. 3 control) and at day 21 after cessation of liraglutide administration (n=3 treated *vs*. 3 control). Images are representative of three mice, and the scale bar represents 25 µm. Data are presented as mean ± SEM. *, *p* <0.05, **, *p* <0.01, ***, *p* <0.001.

Then, risk factors for delayed perfusion were further investigated. Except for a temporary decrease in body weight in the treated group, none of the variables, body weight, blood glucose, or blood pressure, showed differences between the treated and control groups, and the delays were not correlated with any of these factors within the treated group **(Extended Fig. 1b)**. As GLP-1RA has been found to reduce intracranial pressure (ICP) in both animal studies and clinical trials^14-16^ and because changes in ICP can affect the ophthalmic artery as it passes through the subarachnoid space, we compared ICP between the treated and control groups. The GLP-1RA–treated group exhibited a reduction in ICP on day 7 after treatment, when we identified delays in injection-to-retina time and AVTT **(Fig. 3a)**. This reduction subsided by day 21 after treatment **(Fig. 3a)**, when AVTT no longer showed differences. Within the GLP-1RA treated group, there was a trend toward a negative correlation between ICP and injection-to-retina time **(Fig. 3b)**. However, as animals in the control group with similarly low ICP did not show arterial filling delay, changes in ICP may be a better indicator and have a more significant correlation instead. Taken together, the results of decreased ICP and delayed retinal perfusion at corresponding time points, and a possible negative correlation within the treated group imply that low ICP induced by GLP-1RA may contribute to delayed perfusion. The decreased ICP might affect the ophthalmic arteries and veins in the subarachnoid space by narrowing the subarachnoid space (**Fig. 3c)**, a phenomenon that can also be observed as decreased optic nerve sheath diameter in intracranial hypotension^17^.

**Figure 3.**
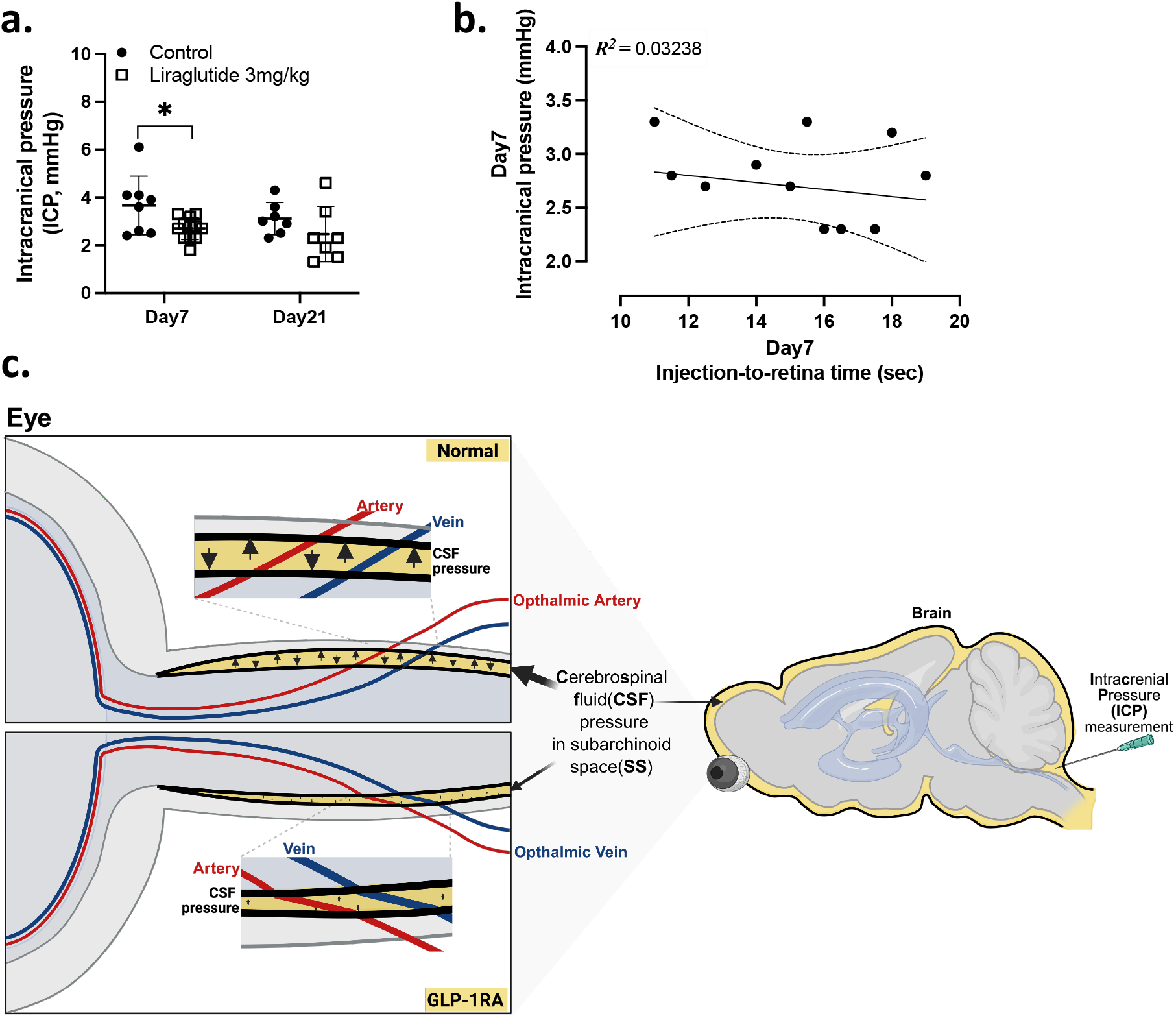
Reduction of intracranial pressure with GLP-1RA liraglutide-treated mice. **a**. Intracranial pressure (ICP) measurement at day 7 (n=8 treated *vs*. 12 control), and at day 21 (n=7 treated *vs*. 7 control) after daily administration of 3 mg/kg liraglutide. **b**. Linear regression between ICP and injection-to-retina time at day 7 after daily liraglutide administration. **c**. Working model of GLP-1RA-induced ICP decrease on retinal perfusion delay. Data are presented as mean ± SEM. *, *p* <0.05. ICP, intracranial pressure; CSF, cerebrospinal fluid.

In this study, we demonstrated delayed retinal perfusion in individuals treated with GLP-1RA, which was also observed in treated mice, accompanied by evidence of retinal ischemia that recovered following treatment discontinuation. Considering the early worsening of diabetic retinopathy induced by GLP-1RA treatment^18,19^, a phenomenon not documented in other circulatory systems^20^, and the rare but clinically significant occurrence of ischemic complications such as NAION^21^ in the absence of systemic ischemic findings, these findings suggest that the unique anatomical and physiological characteristics of ocular circulation may render it particularly vulnerable to GLP-1RA-related effects. Investigation of ICP revealed that treated mice exhibited decreased ICP, which may contribute to the observed delay in retinal perfusion. Further investigation of temporal ICP changes in humans could help elucidate these mechanistic connections. Although our animal model lacks a lamina cribrosa and did not develop any signs of NAION, we consistently demonstrated delayed retinal perfusion across both human and mice. Therefore, careful monitoring of ocular circulation may be warranted in individuals using GLP-1RA, regardless of underlying comorbidities.

## Data Availability

All data produced in the present study are available upon reasonable request to the authors

## Acknowledgements

We would like to express our gratitude to the participants, without whom this research would not have been possible. This study was supported by the National Research Foundation of Korea (NRF) grant funded by the Korea government (MSIT) (RS-2024-00359653 and RS-2024-00512914) and by grant no. 04-2025-0350 from the SNUH Research Fund. The funders had no role in study design, data collection and analysis, decision to publish, or preparation of the manuscript.

## Author Contributions Statement

Y.K.L. designed experiments, collected and analyzed data, and wrote the manuscript. Y.Y.C. performed experiments and collected data. H.N.J. collected and analyzed clinical data. D.J.M. analyzed clinical data. H.B.S. conceptualized, supervised the study, analyzed clinical data, and wrote the manuscript.

## Figure Legends

**Extended Figure 1. Mouse retina optical coherence tomography (OCT) and measurements of body weight, blood glucose, and blood pressure during the liraglutide treatment and post-treatment periods**.

**a**. Mouse retina optical coherence tomography (OCT) measurements at day 7 after daily administration of 3 mg/kg liraglutide (n=6 treated *vs*. 7 control). **b**. Mouse body weight and blood glucose changes during the treatment and post-treatment periods. Mouse blood pressure measurement at day 21 after daily liraglutide administration (n=7 treated *vs*. 8 control). Data are presented as mean ± SEM. *, *p* <0.05, **, *p* <0.01. OCT, optical coherence tomography.

**Extended Table 1.** Baseline characteristics of GLP-1RA prescribed patients in type 2 diabetes.

|  | Mid-GLP-1RA<br>Treatment (n=6) | Post-GLP-1RA<br>Cessation (n=3) | *p-value* |
| --- | --- | --- | --- |
| Age(yrs) | 68 ± 17 | 63 ± 3 | 0.571 |
| Sex (M:F) | 3:3 | 2:1 | 1.000 |
| Height (cm) | 164.92 ± 12.35 | 160.90 ± 14.99 | 0.712 |
| Body weight(kg) | 80.88 ± 23.88 | 80.57 ± 17.59 | 0.983 |
| BMI (kg/m<sup>2</sup>) | 29.18 ± 5.31 | 30.96 ± 4.07 | 0.600 |
| HbA1c (%) before GLP-1RA treatment | 8.53 ± 1.29 | 9.23 ± 3.10 | 0.739 |
| HbA1c (%) after 52wk of GLP-1RA treatment | 7.40 ± 0.91 | 7.60 ± 1.23 | 0.818 |
| Diabetic retinopathy grade<br>(Mild:Moderate:Severe NPDR:PDR) | 1:2::2:1 | 0:2:1:0 | 1.000 |
| FAG interval (wks) | 78.52 ± 33.89 | 136.19 ± 72.42 | 0.299 |
| GLP-1RA prescription (wks) | 197.1 ± 55.68 | 127.1 ± 34.48 | 0.058 |
| Highest dulaglutide dose (mg/ml) | 3 | 3 | 1.000 |

BMI, body mass index; HbA1c, hemoglobin A1c; FAG, fluorescein angiography; GLP-1RA, glucagon like peptide-1 receptor agonist.

**Extended Table 2.**
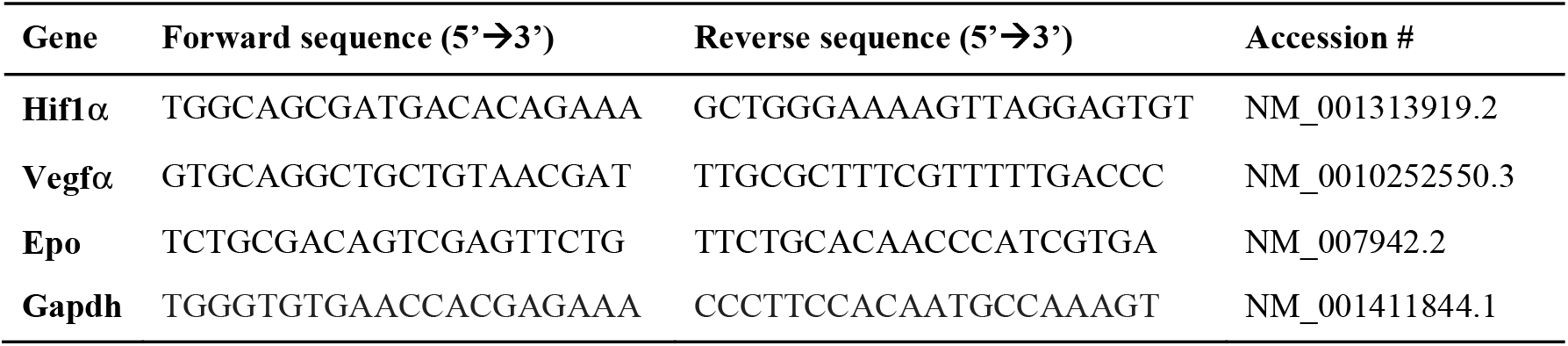
Mouse primer sequence list.

## Methods

### Study patients

The study used data from the Kangbuk Samsung Medical Center, which underwent regular checkups in the outpatient department with fluorescein angiography (FAG) before and after GLP-1RA dulaglutide administration in type 2 diabetes patients. This study was approved by the Institutional Review Board of Kangbuk Samsung Hospital (IRB: KBSMC 2025-04-019), with informed consent waived due to the use of anonymized data.

### Animals

Adult male mice (C57BL/6, 6–7 weeks old) were purchased from ORIENT BIO (Korea) and housed in a temperature-controlled environment with free access to food and water under a 12 h light-dark cycle. Liraglutide (Saxenda, Novo Nordisk, Denmark) was administered intraperitoneally (i.p.) at a dose of 3 mg/kg daily for 21 days. All animal experimental protocols were approved by Seoul National University IACUC (Institutional Animal Care and Use Committees; SNU-240408-1, SNU-250205-1-1, SNU-250122-3-2), and all experiments were carried out following ARVO Statement for the Use of Animals in Ophthalmic and Vision Research and the international standards of animal care and use, ARRIVE (Animal Research: Reporting in Vivo Experiments) guidelines.

### Retinal Perfusion Analysis

Mice were anesthetized with a mixture of Zoletle (40 mg/kg) and Rompun (5 mg/kg) (1:3 mixture) administered by intraperitoneal (i.p.) injection, and 100 μL of FITC (fluorescein isothiocyanate)-dextran (25 mg/mL, 2000 kDa, Sigma, USA) was injected into the mouse through the lateral tail vein with a 30-gauge syringe needle (BD, USA). Then retina was imaged with a phoenix micron IV retinal imaging laser camera (Phoenix Research Labs, USA). Initial perfusion time was evaluated by measuring the time between the injection and the first visualization of retinal vessels. Arteriovenous transit time was assessed by measuring the time between the artery and vein in the retina during retinal perfusion recording. At seven days and twenty-one days during drug treatment, the retina was imaged for perfusion and A-V transition time.

### Electroretinography (ERG)

Mice were dark-adapted for 18 hrs before testing. Under dim red illumination, anesthesia was administered by intraperitoneal injection with a mixture of zoletile (40 mg/kg) and rompun (5 mg/kg) (1:3 mixture). Pupils were dilated with 2.5% phenylephrine hydrochloride and 1% tropicamide (Hanmi, Korea) eye drops, then replaced with a 2% hypromellose solution for ERGs. ERGs were recorded using the combined stimulator/electrode probe of the Celeris system (Diagnosys, LLC, Lowell, USA), which was placed in contact with each cornea and aligned with the center of the pupils. Under this setup, stimuli were presented to one eye at a time along with the other eye. Data obtained from the left and right eyes were averaged for analysis. Recording was performed under heating pad. Amplifier bandpass settings were 0.01 to 10 Hz.

### Optical Coherence Tomography (OCT)

Optical coherence tomography was performed with IISCIENCE OCT for all animals after 7 days of treatment (iiScience, USA). Mice were anaesthetized with a mixture of Zoletil (40 mg/kg) and Rompun (5 mg/kg) (1:3 mixture) administered by intraperitoneal (i.p.) injection. To take imaging, pupils were dilated with 2.5% phenylephrine hydrochloride and 1% tropicamide (Hanmi Pharm, Korea), then the dilated solution was removed and replaced with a 2% Hypromellose solution during image acquisition (Samil Pharm, Korea). For OCT image acquisition, the camera was positioned close to the eye until the pupil was in focus on the live frame. At this point, the camera head was fixed, and by using the reference arm slider, the cornea and the lens reflection image were positioned at the optimal spot for the acquisition window.

### Anesthesia and Surgical Procedures

To measure ICP, the mice were treated for 7 days and then anesthetized with a mixture of Zoletil (40 mg/kg) and Rompun (5 mg/kg) (1:3 ratio), administered intraperitoneally (i.p.) injection. Afterwards, the head skin was sterilized and incised down to the skull. The head and upper body are positioned at approximately a 90° angle, and the space between the skull and the first cervical vertebra where the needle is inserted is opened, aiming into the cisterna magna. The needle’s bevel, connected to a pressure manometer, was gradually penetrated and faced upward into the cisterna magna (Kovax syringe 26G1/2”, Korea; Benetech GW510, China). Measurement was performed for about 5 minutes until the pressure started to decline.

### Blood Pressure Measurements

Systolic and diastolic blood pressure were measured by tail-cuff plethysmography using a BP-2000 blood pressure analysis system (Visitech Systems, Apex, NC, USA). Mice were placed in a metal box restraint with the tail passing through the optical sensor and compression cuff before finally being taped to the platform. A traditional tail-cuff occlude was placed proximally on the animal’s tail, which was then immobilized with tape in a V-shaped block between a light source above and a photoresistor below. Upon inflation, the occlude stopped blood flow through the tail, while upon deflation, the sensor detected the return of blood flow. The restraint platform was maintained at a temperature of 37 °C. Before experiments, mice were acclimated to restraint and tail-cuff inflation for 2 days. On the test day, 10 measurements were made to collect basal blood pressure.

### Immunohistochemistry

Eyeballs were enucleated from the mouse and then fixed in 4% PFA. Eyeballs were removed from the fixative, and an incision was made on the cornea. The eyeball was transferred to 10% sucrose in PBS for 1 hour, then transferred to 20% sucrose in PBS for 1 hour, and lastly transferred to 30% sucrose in PBS overnight at 4 °C for cryosection. The eyecup was embedded into the optimal cutting temperature (OCT) compound with a cryomold. After it was snap-frozen with isopentane in liquid nitrogen, the tissue was sectioned on a cryostat at 10 µm. The retina sections were permeabilized using 5% donkey serum in PBS containing 0.5% Triton X-100. The sections were incubated overnight at 4 °C with the primary antibody anti-Hif1α (1:400, Cell Signaling #36169, USA). The sections were then detected with species-appropriate fluorescence-conjugated secondary antibodies (Thermo Fisher Scientific, USA). The retina image was acquired and analyzed with a FV3000 confocal laser scanning microscope and examined using the FV31S-SW Viewer (FV3000, Olympus, Japan).

### RNA extraction, cDNA synthesis, and quantitative real-time PCR

Total RNA was extracted from the isolated mouse retina using TRIzol reagent (Invitrogen, USA). Reverse transcription of 1 μg RNA was performed with a high-capacity cDNA reverse transcription kit (Thermo Fisher Scientific, USA). Quantitative real-time PCR was conducted using SYBR Green PCR Master Mix (Enzynomics, Korea) and specific primers (listed in **Extended Table 2**). Reactions were carried out using a QuantStudio 6 Flex Real-Time PCR System (Life Technology, USA). Relative mRNA levels were normalized to actin/GAPDH expression and expressed as a fold change relative to controls.

### Statistical Analysis

All experiments were repeated at least 3 times, and data analyses were performed with GraphPad Prism 10 (USA). Data are presented as mean ± SEM. Time-dependent differences were analyzed using two-way repeated-measures ANOVA with Bonferroni post hoc tests. Comparisons between groups were performed using the Mann–Whitney U test or Kruskal-Wallis test with Dunn’s multiple comparisons. Statistical significance was set at *p* < 0.05

