## Supplemental Table1, Supplemental Table2 for "Retinal perfusion delay associated with GLP-1 receptor agonists: A possible role of intracranial pressure?"

### Extended Table 1.

| | During-GLP-1RA Treatment (n=6) | Post-GLP-1RA Cessation (n=3) | *p-value* |
|---|---|---|---|
| **Age (yrs)** | 68 ± 17 | 63 ± 3 | 0.571 |
| **Sex (M:F)** | 3:3 | 2:1 | 1.000 |
| **Height (cm)** | 164.92 ± 12.35 | 160.90 ± 14.99 | 0.712 |
| **Body weight(kg)** | 80.88 ± 23.88 | 80.57 ± 17.59 | 0.983 |
| **BMI (kg/m$^2$)** | 29.18 ± 5.31 | 30.96 ± 4.07 | 0.600 |
| **HbA1c (%) before GLP-1RA treatment** | 8.53 ± 1.29 | 9.23 ± 3.10 | 0.739 |
| **HbA1c (%) after 52wks of GLP-1RA treatment** | 7.40 ± 0.91 | 7.60 ± 1.23 | 0.818 |
| **Diabetic retinopathy grading (Mild:Moderate:Severe NPDR:PDR)** | 1:2:2:1 | 0:2:1:0 | 1.000 |
| **FAG interval (wks)** | 78.52 ± 33.89 | 136.19 ± 72.42 | 0.299 |
| **GLP-1RA prescription (wks)** | 197.1 ± 55.68 | 127.1 ± 34.48 | 0.058 |
| **Highest dulaglutide dose (mg/ml)** | 3 | 3 | 1.000 |

BMI, body mass index; HbA1c, hemoglobin A1c; FAG, fluorescein angiography; GLP-1RA, glucagon like peptide-1 receptor agonist.

### Extended Table 2.

| Gene | Forward sequence (5'→3') | Reverse sequence (5'→3') | Accession # |
|---|---|---|---|
| **Hif1α** | TGGCAGCGATGACACAGAAA | GCTGGGAAAAGTTAGGAGTGT | NM_001313919.2 |
| **Vegfα** | GTGCAGGCTGCTGTAACGAT | TTGCGCTTTCGTTTTTGACCC | NM_0010252550.3 |
| **Epo** | TCTGCGACAGTCGAGTTCTG | TTCTGCACAACCCATCGTGA | NM_007942.2 |
| **Gapdh** | TGGGTGTGAACCACGAGAAA | CCCTTCCACAATGCCAAAGT | NM_001411844.1 |
