## Supplementary figures and images for "Retinal perfusion delay associated with GLP-1 receptor agonists: A possible role of intracranial pressure?"

### Supplemental Figure 1

#### Extended Figure 1.

**a.**

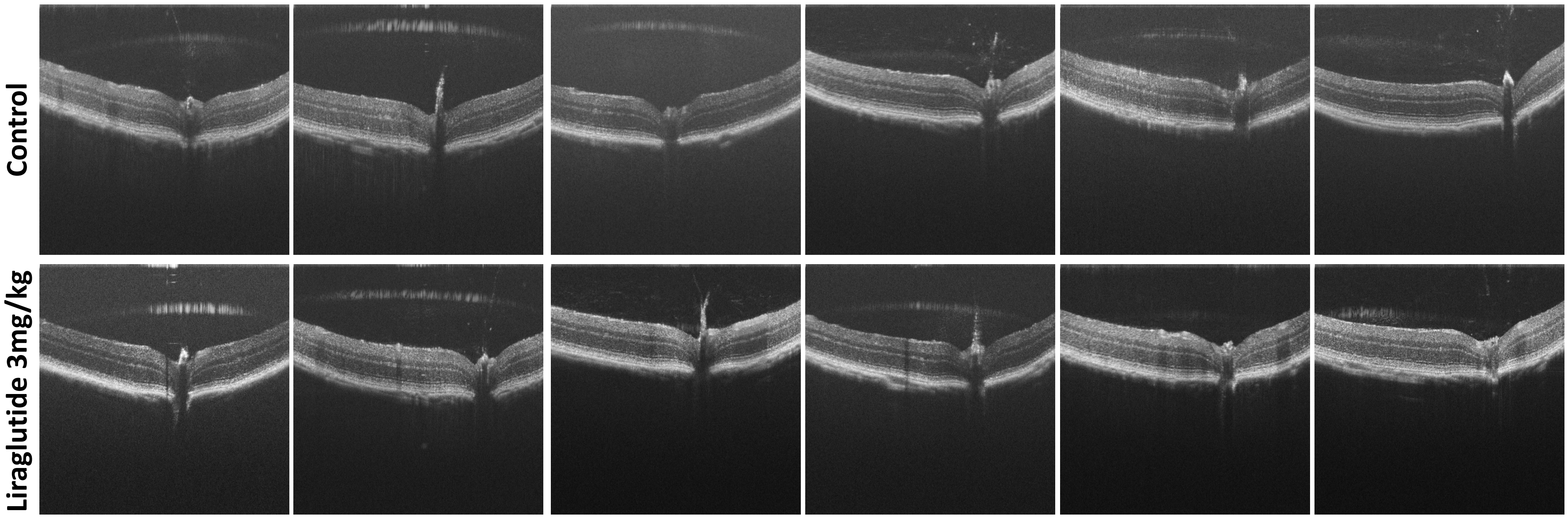


**b.**

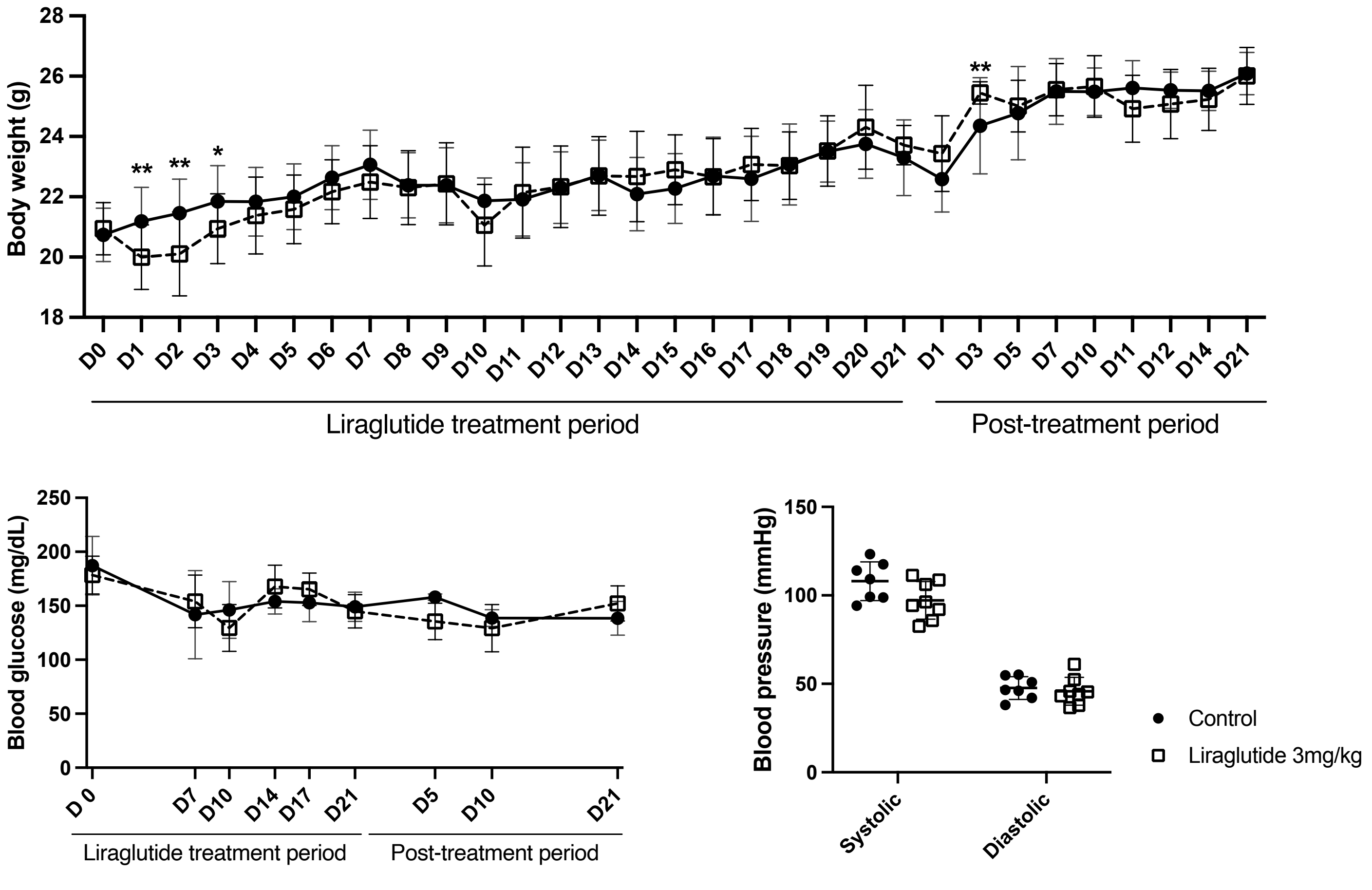
